# Acute Dose-Response Effects of Guarana and Caffeine on Cognitive Performance in Healthy Adults

**DOI:** 10.64898/2026.11.12.26367967

**Authors:** Ziyue Huang, Tae Lee, Tom Gurney, Terun Desai

## Abstract

Guarana (*Paullinia cupana*) is a fruit native to the Brazilian Amazon. It exerts potential cognitive-enhancing effects, primarily attributed to its caffeine content and additional bioactive compounds. However, the dose-response relationship of guarana and its comparative effects on cognitive performance to caffeine remain unclear. This double-blind, placebo-controlled, randomised crossover study aimed to investigate the acute effects of guarana at three different doses (containing 1, 2, and 4 mg⋅kg⋅bodyweight^-1^ caffeine) compared to matched-dose caffeine (4 mg⋅kg⋅bw^-1^) and placebo on cognitive performance. Nineteen healthy adults completed one baseline assessment and five experimental sessions on six different days, each one week apart. During each experimental session, participants consumed one of the five supplement drinks in a randomised order. Four different cognitive tasks (memory, simple reaction time, sustained attention, and inhibition tasks) were performed pre-ingestion and 30 minutes post-ingestion to assess cognitive performance.

Reaction time and number of errors in tasks were recorded, and data were analysed using a repeated measures analysis of variance. A significant main effect of session was found for memory (*p* = 0.05) and sustained attention (*p* = 0.01). However, no significant main effects of treatments, times, or treatment-by-time interactions were observed on reaction time or number of errors in all variables (*p* > 0.05). Guarana ingestion was not significantly different to matched-dose caffeine or placebo, and no dose-response relationship was observed for memory, simple reaction time, sustained attention, and inhibition tasks. Further research is warranted to explore chronic use, individual variability, and underlying physiological mechanism of guarana.

## 1. Introduction

Guarana (*Paullinia cupana*), a native fruit of the Brazilian Amazon, has been widely recognised for its stimulant and medicinal properties due to its rich composition of bioactive compounds, including caffeine, tannins, saponins, flavonoids (e.g., catechins, epicatechins) and other methylxanthines (e.g., theobromine, theophylline) (1). Guarana was historically used by indigenous populations for its energising effects (2), and it is now commonly included in energy drinks and dietary supplements (1,2). In addition to its stimulant effects, guarana exhibits antioxidant, anti-inflammatory and diuretic functions (1,3). Emerging evidence also highlights potential clinical applications, such as reducing fatigue in cancer patients (4), improving metabolic health in older populations (5), and aiding short-term weight loss (6).

Guarana’s potential cognitive benefits have also gained increasing attention, primarily due to its exceptionally high caffeine content (up to 6% of its dry weight, compared to about 2% in coffee beans) (2). Caffeine, a well-established cognitive enhancer (7), is the primary bioactive compound in guarana (2). It crosses the blood-brain barrier and acts as an adenosine receptor antagonist (8), leading to increased release of neurotransmitters such as norepinephrine and dopamine (9), which improve alertness and attention (7). Caffeine is primarily metabolised in the liver via cytochrome P450 enzymes, with cytochrome P450 1A2 (CYP1A2) being the main isoenzyme responsible (10). CYP1A2 catalyses N-3 demethylation of caffeine to produce paraxanthine, accounting for 90% of caffeine metabolism (11).

Cognitive performance encompasses interrelated domains including attention, memory and executive control, all of which are integral to academic and occupational success, yet sensitive to arousal and mental fatigue (12,13). Several studies have explored guarana’s influence on these domains, with findings suggesting improvements in reaction time, working memory, information processing, and subjective alertness (14–21). For example, Gurney et al. (14) observed reductions in simple and choice reaction times (RTs) following guarana supplementation. Similarly, Pomportes et al. (17) noted quicker information processing (decreased time taken) with higher accuracy following guarana supplementation via serial mouth rinsing. However, the literature remains inconsistent, with some investigations reporting null effects on reaction time or task accuracy (14,15,18,22). Veasey et al. (18) reported improved numeric working memory accuracy, whereas Kennedy et al. (15) observed no changes in attentional task accuracy with guarana. Such variability may reflect differences in guarana dose, participant characteristics, and cognitive task sensitivity (17,18,23). Importantly, beneficial effects of guarana supplementation have been observed at doses containing caffeine levels below the recognised threshold for psychostimulant activity (0.5 mg⋅kg⋅bw^-1^) (7,15), suggesting that guarana’s non-caffeine constituents may contribute to beneficial cognitive outcomes. Flavonoids, for example, possess antioxidant properties that might mitigate oxidative stress in the brain and support cognitive function (24), while theobromine has been shown to act synergistically with caffeine to improve aspects of mood and cognition (25). Supporting this, several studies indicate that guarana produces stronger cognitive effects than equivalent doses of caffeine alone (14,17,23). Conversely, different trials from the same studies also report no meaningful differences between guarana and caffeine, evident to task-specific demands or methodological variability (14,17,23).

A further limitation of the current evidence lies in the uncertainty surrounding optimal dosing. Reported guarana doses ranges widely from 1 – 125 mg⋅kg⋅bw^-1^, with reported caffeine content (0.12 – 5 mg⋅kg⋅bw^-1^) varied across different experiments (22). To date, only two studies have assessed the dose-dependent effects of guarana on cognitive performance. One study reported greater memory improvements at lower dose (37.5 mg) than higher dose (300mg) (16), while meta-analytic findings indicated no clear dose-response relationship (22).

Therefore, this research aimed to determine and compare the acute effects of different doses of guarana (containing 4 mg⋅kg⋅bw^-1^, 2 mg⋅kg⋅bw^-1^, or 1 mg⋅kg⋅bw^-1^ caffeine) and caffeine (4 mg⋅kg⋅bw^-1^) on cognitive performance, including alertness, concentration, inhibitory control, and memory in healthy adults. The cognitive performance was assessed via RT and number of errors in simple reaction time (SRT), sustained attention (SA), inhibition (INH), and memory (MEM) tasks before and after supplementation. It was hypothesised that the highest guarana dose (containing 4 mg⋅kg⋅bw^-1^caffeine) would produce the greatest improvement in RT and the largest reduction in errors in all tasks, compared to the matched dose of caffeine and lower guarana doses used in this study.

## 2. Methods

### 2.1 Participants

Twenty-three healthy adults were initially recruited through convenience sampling. Four participants voluntarily withdrew from the study due to personal reasons unrelated to the study procedures. As a result, the final analysed sample consisted of nineteen healthy adults; 13 males and 6 females as shown in *Table 1*.

**Table 1.** Participant Characteristics presented as mean ± SD for the final cohort completing the study (n=19).

|  | All (N=19) |  | Female (N=6) |  | Male (N=13) |  |
|---|---|---|---|---|---|---|
|  | Mean | SD | Mean | SD | Mean | SD |
| Age (years) | 22 | 4 | 23 | 4 | 22 | 5 |
| Stature (m) | 1.8 | 8.5 | 1.7 | 5.1 | 1.8 | 5.5 |
| Body Mass (kg) | 71.0 | 10.6 | 62.2 | 9.3 | 74.4 | 9.4 |
| BMI (kg·m<sup>-2</sup>) | 22.7 | 2.7 | 22.7 | 3.8 | 22.8 | 2.4 |
| Body Fat Percentage (%) | 16.5 | 6.8 | 24.7 | 5.8 | 13.3 | 3.8 |
| Muscle Mass (kg) | 56.5 | 10.0 | 44.4 | 7.1 | 61.1 | 6.4 |

*Note: No statistically significant gender differences were observed in participant characteristics (p > 0.05).*

Recruited participants were healthy individuals aged above 18 years. Exclusion criteria included known intolerance to caffeine, guarana, Acesulfame K, or Sucralose; cardiovascular conditions or exercise contraindications identified via the Physical Activity Readiness Questionnaire (PAR-Q); pregnancy or suspected pregnancy; and regular smoking or vaping. All participants provided written informed consent and completed PAR-Q before participation. Ethical approval was granted by the UCL Research Ethics Committee (ID: 21745/004) in line with the Declaration of Helsinki.

A priori power analysis (*α* = 0.05, 1 - *β* = 0.80, η² = 0.25 (26)) indicated that a minimum sample of 17 was required to detect statistically significant effects of guarana on cognitive performance by analysing reaction times and number of errors using repeated-measures within-factors ANOVA.

### 2.2 Study design

The study employed a double-blind, placebo-controlled, randomised crossover design. Participants attended the laboratory on 6 separate days, each one week apart. All sessions were conducted at the same time of day in a controlled 19°C environment (14,23). Visit 1 consisted of a familiarisation of the cognitive testing battery (9,27–29). Visits 2-6 served as experimental trials with cognitive tests. Before each visit, participants completed a 24-hour food log and refrained from vigorous exercise, caffeine and alcohol for 24 hours. At the beginning of each visit, participant’s stature (seca 704, seca, Germany) and body mass (seca 704, seca, Germany) were measured. A schematic protocol is shown in *Figure 1*. For the five experimental visits, individuals completed a pre-supplement cognitive battery, consumed their assigned supplement within two minutes, and rested for 30 minutes to allow absorption (14,18,30). During this rest period, participants remained seated in a quiet testing area to minimise external stimulation or physical exertion. This duration was chosen based on evidence that caffeine typically reaches peak plasma concentration within 30-60 minutes (30), while guarana’s cognitive effects have also been observed within a similar timeframe (14,18). The same cognitive battery was then repeated after 30 minutes to assess post-supplementary effects.

**Figure 1.**
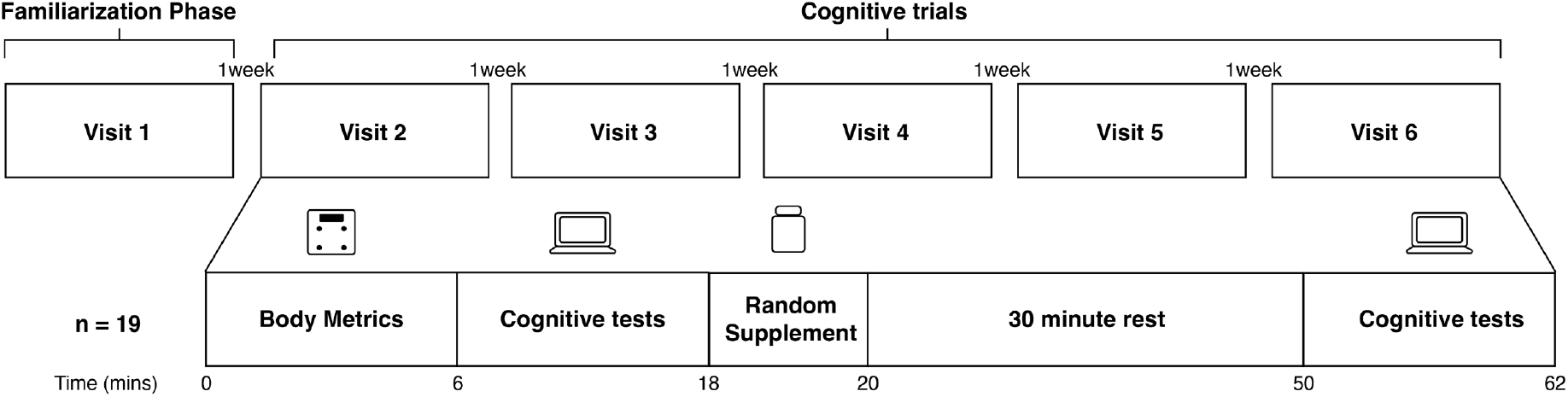
Schematic diagram of the six-visit experimental protocol across six weeks. Assessments included anthropometric measurements and cognitive tests (memory, simple reaction time, sustained attention, and inhibition). The randomised supplements could be a 250 mL drink containing either placebo, caffeine, or guarana with caffeine doses of 4 mg⋅kg⋅bodyweight (bw)^-1^, 2 mg⋅kg⋅bw^-1^, or 1 mg⋅kg⋅bw^-1^.

An exit questionnaire was administered after Visit 6 to evaluate blinding efficacy which showed a 30% correct identification rate.

### 2.3 Supplementation

During visits 2-6, participants received one of the five drinks in a randomised order: caffeine (4 mg⋅kg⋅bw^-1^) (Oxford Vitality, Bicester), guarana (Seven Hills Wholefoods, Sheffield) containing 4 mg⋅kg⋅bw^-1^, 2 mg⋅kg⋅bw^-1^, 1 mg⋅kg⋅bw^-1^ caffeine, or placebo (150 mL water and 100 mL orange squash (Lindhouse, Govone)). The guarana’s caffeine content was verified using high-performance liquid chromatography and reported to be 3.37%. The 4 mg⋅kg⋅bw^-1^ caffeine dose was selected based on its proven efficacy for improving cognitive and physical performance (7) and following International Olympic Committee guidelines (31).

Supplements were mixed in 150 mL room-temperature water and 100 mL squash to standardise taste and texture. An independent staff member prepared and dispensed drinks in opaque bottles to ensure blinding of participants and researchers. Participants were instructed to pinch their noses during consumption to minimise olfactory cues.

### 2.4 Dietary control

Participants were told to maintain their habitual diet, avoid consumption of other supplements throughout the study, and abstain from caffeine consumption 24 hours before testing sessions. This approach ensured guarana’s polyphenols complemented habitual polyphenol intake, reflecting typical consumption and enhancing real-world relevance.

Participants’ twenty-four-hour food diaries prior to each visit reflected a typical Western diet and were analysed with Dietplan 7.0 (Forestfield Software, UK) to assess energy intake, macronutrients, polyphenols, and caffeine. This analysis established baseline dietary intake and ensured dietary intake was similar before each test. The analysis revealed that participants’ macronutrient contributions to total energy intake were as follows: protein (22% ± 9%), carbohydrates (41% ± 22%), fat (37% ± 17%), habitual polyphenol intake (323 ± 333 mg⋅day ⁻¹), and habitual caffeine intake (60 ± 64 mg⋅day ⁻¹). Three of the twenty participants were regular caffeine consumers at 100-300 mg⋅day⁻¹. Habitual caffeine intake across all participants ranged from 0-250 mg.

### 2.5 Cognitive Tasks

Both pre- and post-supplementation cognitive assessments comprised the same four tasks: MEM, SRT, SA, and INH. All tasks were designed and administered using the online Gorilla Experiment Builder (www.gorilla.sc). All tests were conducted in a quiet room without any other individual present, with participants seated 1m from the screen and hands placed on response keys.

#### 2.5.1 The Smiley Task Battery

A smiley task battery was used to assess SRT, SA, and INH (26). The test battery adopted black-and-white ideograms to minimise potential confounding effects from non-executive processing associated with individual differences (26). The tasks were presented in a fixed order: SRT, SA, and INH to optimise construct validity (26). This sequencing may cause the SA task to feel slower and more monotonous following the SRT task, thereby increasing attentional demands (26). Presenting the SRT and SA tasks before the INH task established a routine response to target stimuli, which must then be suppressed during the Inhibition condition, thereby heightening the inhibitory demands of the task (26).

The smiley task’s test-retest reliability was assessed by Ronca et al. (26) in 256 external participants, showing high reliability for RT in SRT (ICC = 0.67), SA tasks (ICC = 0.75), and INH (ICC = 0.76) but lower reliability for number of errors in SA (ICC = 0.28) and INH (ICC = 0.52) (26).

##### 2.5.1.1 Simple Reaction Time Task

The SRT task was used to assess alertness (32–34). Participants were instructed to press the spacebar as quickly as possible when a smiley or winky face appeared on the screen (*Figure 2*). Following 8 practice trials, 30 test trials (15 each) were presented with inter-trial intervals (ITIs) of 1000, 1500 or 2000 ms in a pseudorandomised order. Mean RT was calculated from all correct responses, excluding those faster than 125 ms (35). Incorrect spacebar presses during ITIs were recorded as errors (36).

**Figure 2.**
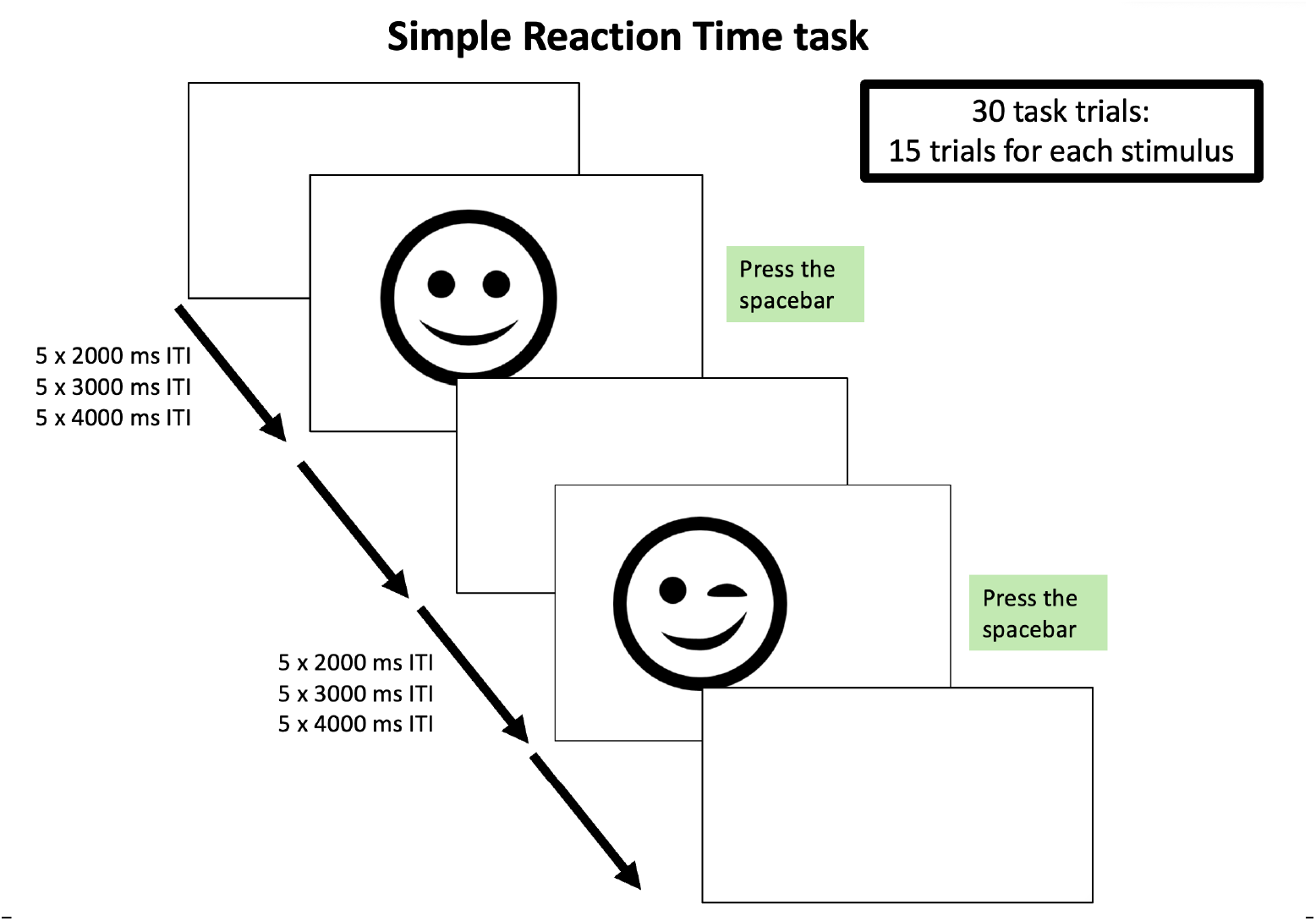
Overview of the simple reaction time (SRT) task with the illustration of visual stimuli used in the task. The number of trials using Smiley and Winky faces, and the number of specific inter-trial intervals were shown.

##### 2.5.1.2 Sustained Attention Task

The SA task adopted a Stay-Go paradigm (*Figure 3*) to assess concentration (37–39). Participants were instructed to press the spacebar in response to a winky face (Go trial) and to withhold response to a smiley face (Stay trial). After 8 practice trials, 30 test trials (10 Go trials were distributed among 20 Stay trials) were administered with ITIs of 2000, 3000 or 4000 ms. Trial types and ITIs were presented in a pseudorandomised order. Mean RT was calculated from correct Go responses, excluding those under 125 ms (35). Errors included omission errors (failures to respond during Go trials) and commission errors (responses during Stay trials) (40).

**Figure 3.**
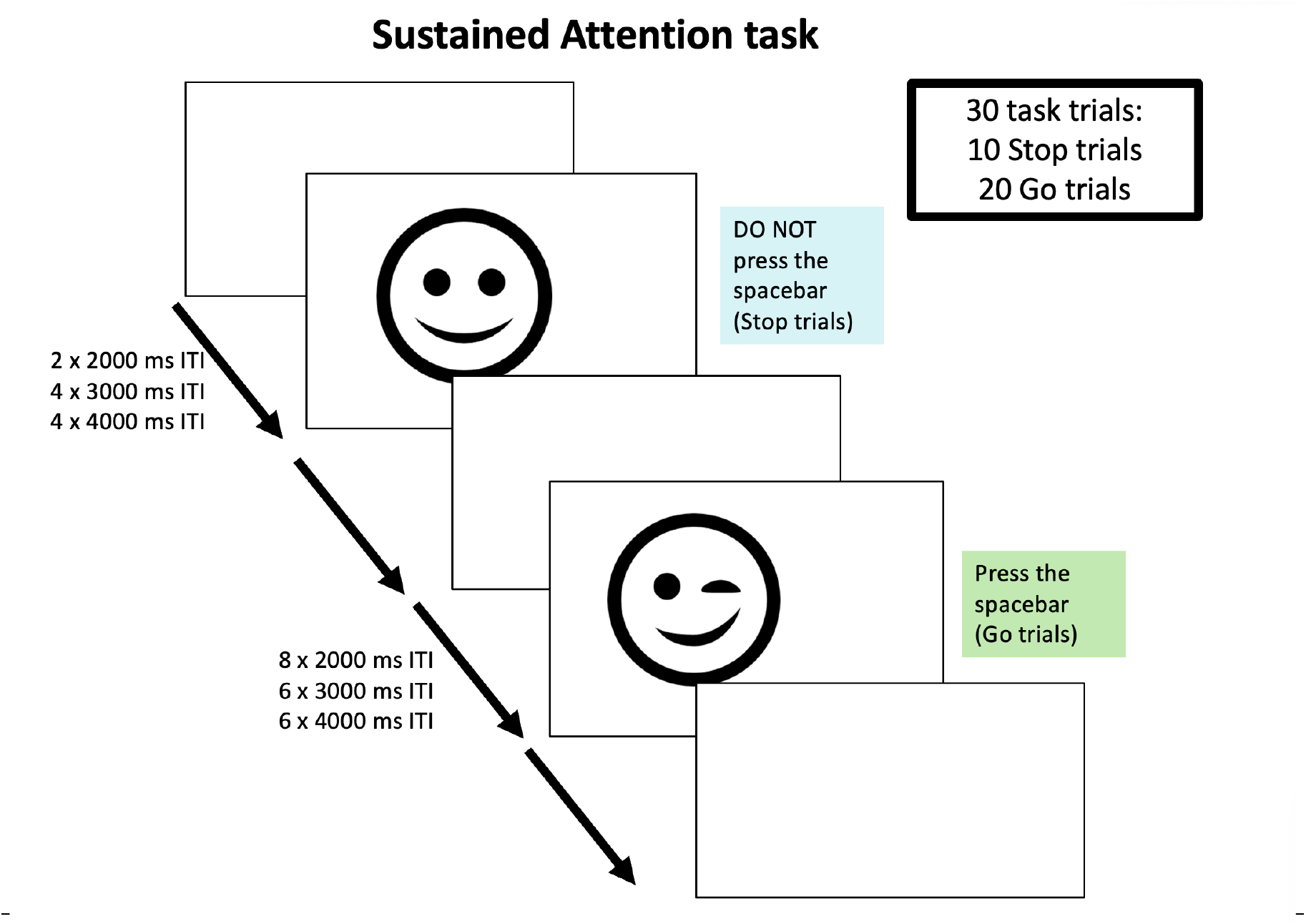
Overview of the sustained attention (SA) task with the illustration of visual stimuli used in the task. The number of trials using Smiley and Winkey faces, and the number of specific inter-trial intervals were shown.

##### 2.5.1.3 Inhibitory Control Task

The INH task employed a Go/No-Go paradigm (*Figure 4*) to measure inhibitory control (41,42). Participants were required to respond to a smiley face (Go trial) by pressing the spacebar and withhold the responses to the winky face (No-Go trial). Eight No-go trials were interspaced with 62 Go trials (500 ms inter-trial interval). Trial types and ITIs were presented in a fixed but pseudorandomised order. Mean RT was calculated from correct Go responses, excluding those under 125 ms (35). Errors included omission errors (failures to respond during Go trials) and commission errors (responses during No-Go trials).

**Figure 4.**
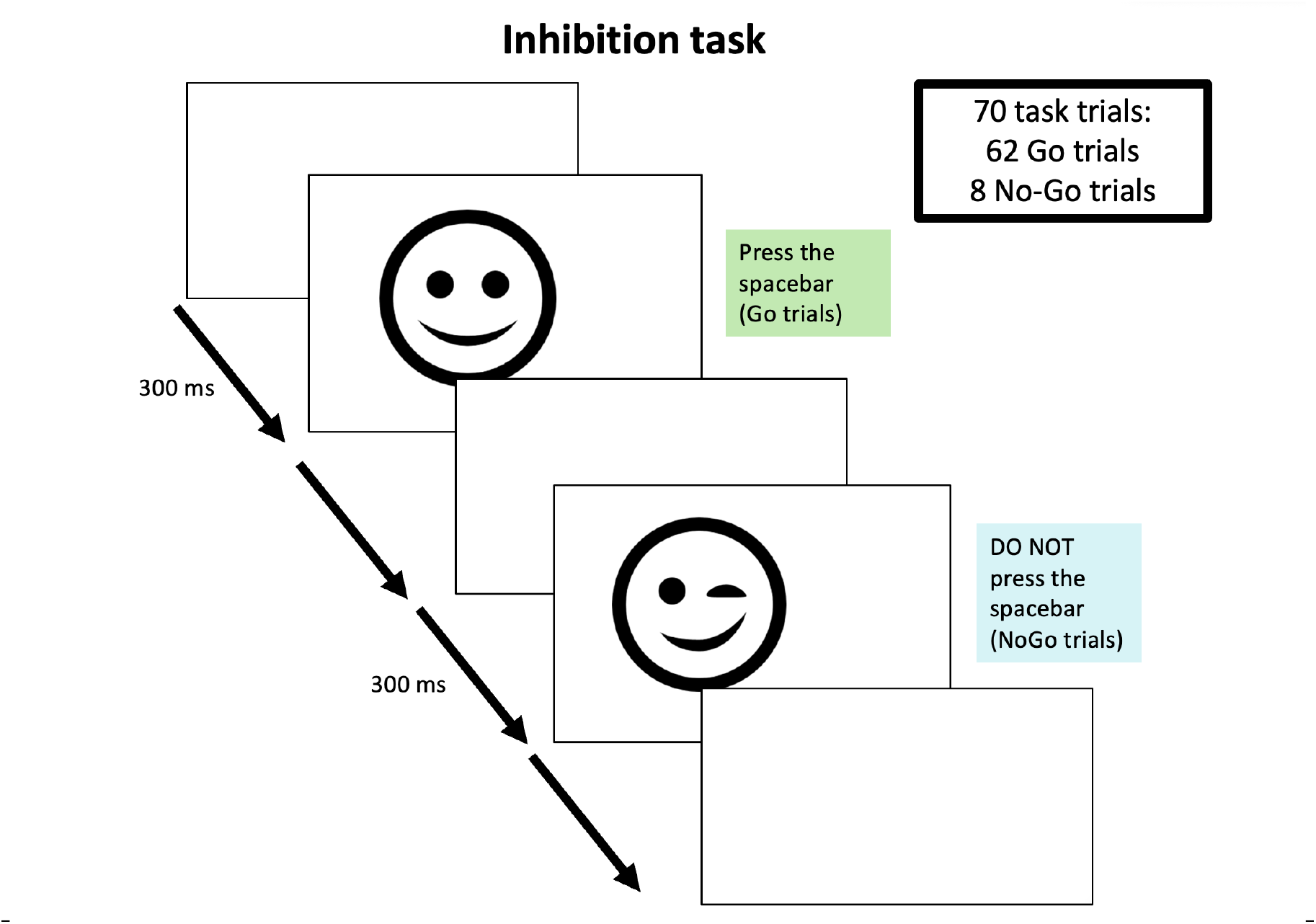
Overview of the inhibition (INH) task with the illustration of visual stimuli used in the task. The number of trials using Smiley and Winkey faces, and the number of specific inter-trial intervals were shown.

#### 2.5.2 The Memory Task

The MEM Task (*Figure 5*) assessed short-term memory using an old/new recognition test (23). During the encoding phase, participants needed to memorise 7 words presented on the screen in a pseudorandomised order. Each word was shown for 1,000 ms. During the retrieval phase, participants received additional instructions to use the mouse to respond to newly randomised word presentations, clicking “Old” if they recognised the word from the earlier phase or “New” if the word was unfamiliar. In total, 17 words (10 new words and 7 old words) were generated for the protocol, which were presented in a pseudorandomised order. The outcome measure was taken as mean reaction time (ms) on correct answers, and the number of errors was defined as the number of incorrect responses recorded.

**Figure 5.**
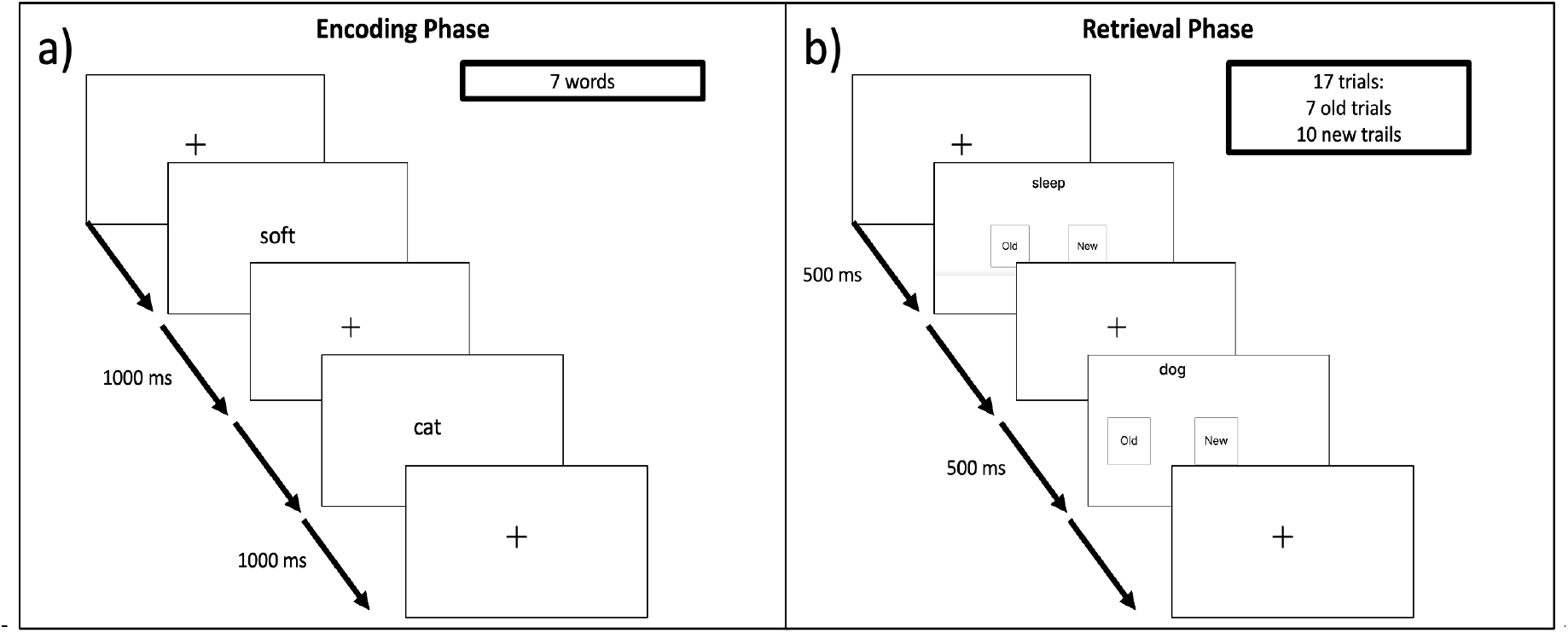
Overview of the memory (MEM) task. **The illustration of visual presentation of the two-phase memory task that consists of a) Encoding phase and b) Retrieval phase.**

### 2.6 Data analysis

Statistical analyses were performed using RStudio (version 2024.12.0+467), with statistical significance set at *p* < 0.05. Data normality was assessed using the Shapiro–Wilk test.

Levene’s test was used to evaluate the homogeneity of variances across times (pre- and post-supplementation) and treatments (placebo, caffeine, and 3 guarana doses). Mean RT and number of errors in SRT and SA tasks were analysed using a 2-way, time (pre-, post-supplementation) × treatments (placebo, 4 mg⋅kg⋅bw^-1^ caffeine, guarana doses containing 1 mg⋅kg⋅bw^-1^, 2 mg⋅kg⋅bw^-1^, and 4 mg⋅kg⋅bw^-1^ caffeine) repeated-measures ANOVA with post-hoc Bonferroni’s adjustment via ‘afex’ R package. When Mauchly’s test indicated a violation of sphericity, the Greenhouse–Geisser correction was applied. Effect sizes were reported using partial eta-squared (η²), with values interpreted as small (0.01 – 0.08), moderate (0.09 – 0.25), large (> 0.25) (43). Data visualisations were generated using ‘ggplot2’ R package. Multiple linear regressions were performed using ‘lm4’ R package to examine treatment effects on RT changes in the SRT and SA tasks, adjusting for age, gender, body fat percentage, and habitual caffeine intake. The model structure was

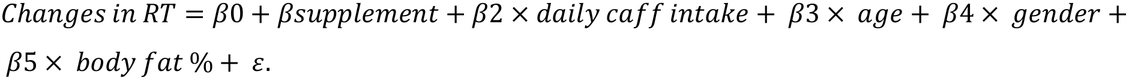

## 3. Results

### 3.1 Simple Reaction Time Task Performance

The mean RTs (*Supplementary Material 1*) in the SRT task were not significantly different among treatments (*F*(_4,72_) = 0.97, *p* = 0.43, η² = 0.01), time (*F*(_1,18_) = 0.62, *p* = 0.44, η² < 0.01), or treatment×time interaction (*F*(_4,72_) = 0.707, *p* = 0.589, η² = 0.02). No main effects for treatments (*F*(_4,72_) = 0.31, *p* = 0.87, η² < 0.01), time (*F*(_1,18_) = 0.40, *p* = 0.54, η² < 0.01), or treatment×time interaction (*F*(_4,72_) = 0.74, *p* = 0.54, η² = 0.01) were detected for number of errors (*Supplementary Material 1*) (*Figure 6*).

**Figure 6.**
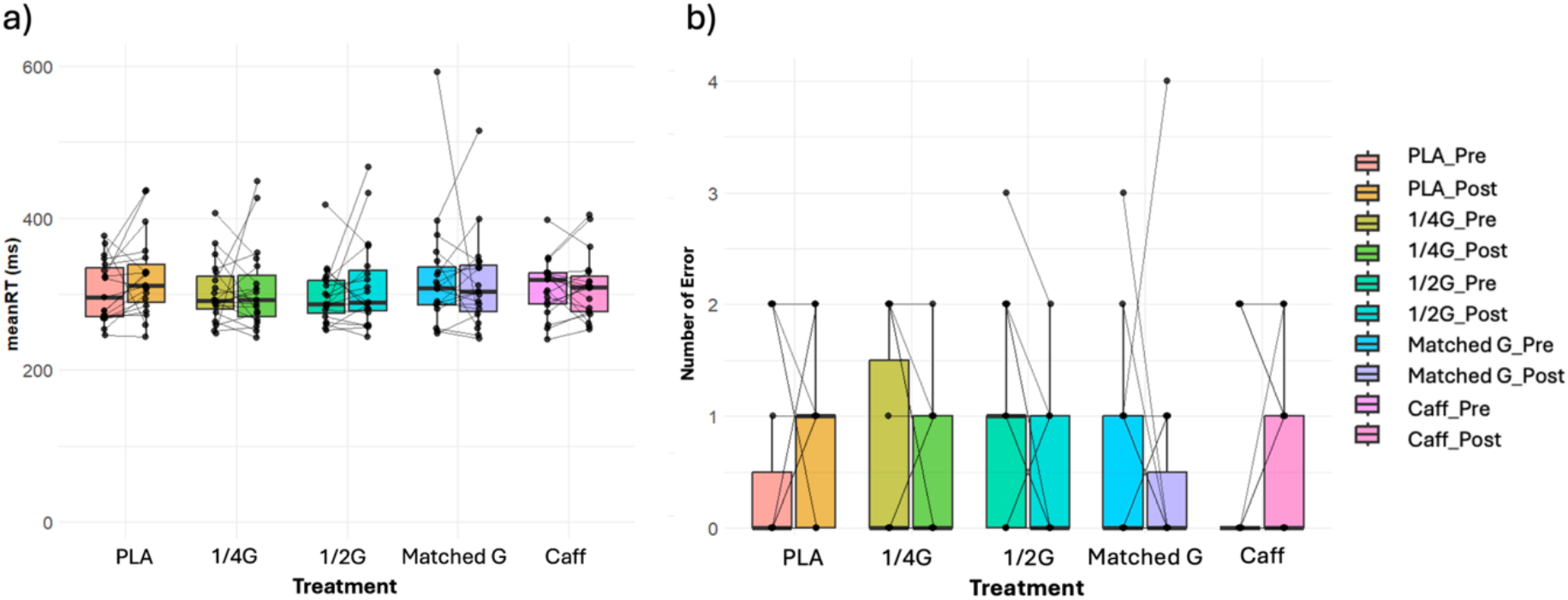
Effects of treatments on SRT task performance with a) Mean Reaction Time (RT) and b) Number of Errors. Dots represent individual performance pre- and post-supplementation; lines connect repeated measures for each participant. RT = reaction time; PLA = placebo; Caff = caffeine; 1/4G, 1/2G, and Matched G represent guarana doses containing 1 mg⋅kg⋅bw^-1^, 2 mg⋅kg⋅bw^-1^, and 4 mg⋅kg⋅bw^-1^ caffeine.

### 3.2 Sustained Attention Task Performance

Individual responses in the SA task across times and treatments are presented in *Figure 7*, highlighting notable variability.

**Figure 7.**
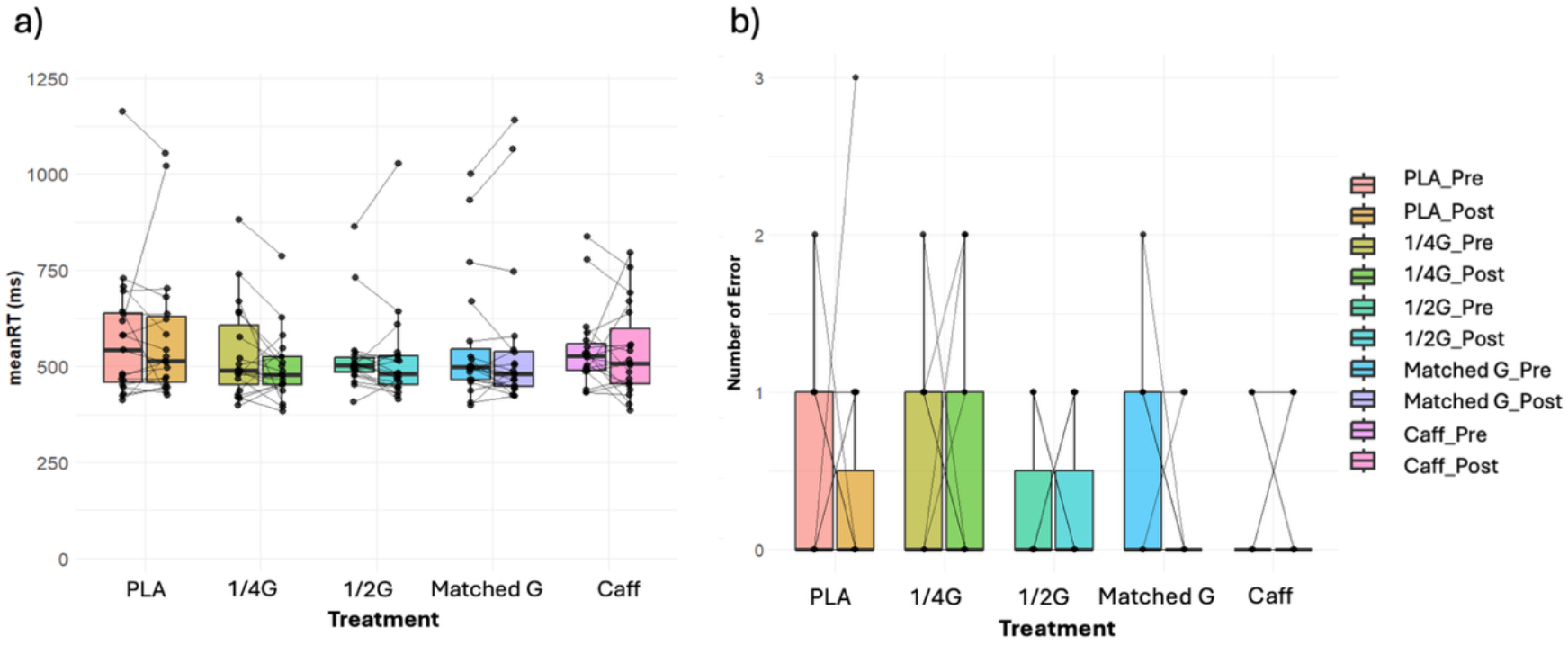
Effects of treatments on SA task performance with a) Mean Reaction Time and b) Number of Errors. Dots represent individual performance pre- and post-supplementation; lines connect repeated measures for each participant. RT = reaction time; PLA = placebo; Caff = caffeine; 1/4G, 1/2G, and Matched G represent guarana doses containing 1 mg⋅kg⋅bw^-1^, 2 mg⋅kg⋅bw^-1^, and 4 mg⋅kg⋅bw^-1^ caffeine.

No statistically significant differences among treatments (*F*(_4,72_) = 1.29, *p* = 0.28, η² = 0.02), time (*F*(_1,18_) = 0.55, *p* = 0.47, η² < 0.01), or treatment×time interaction (*F*(_4,72_) = 0.64, *p* = 0.60, η² < 0.01) were found for mean RT (*Supplementary Material 1*) in the SA task. There were no main effects for treatments (*F*_(4,72)_ = 1.81, *p* = 0.14, η² = 0.03), time (*F*_(1,18)_ = 1.38, *p* = 0.26, η² = 0.01), or treatment×time interaction (*F*_(4,72)_ = 0.78, *p* = 0.54, η² = 0.01) for number of errors (*Supplementary Material 1*).

### 3.3 Inhibition Task Performance

Participant-level data is shown in *Figure 8* for the Inhibition Task assessment across treatments and time, revealing diverse response patterns and substantial between-subject variability.

**Figure 8.**
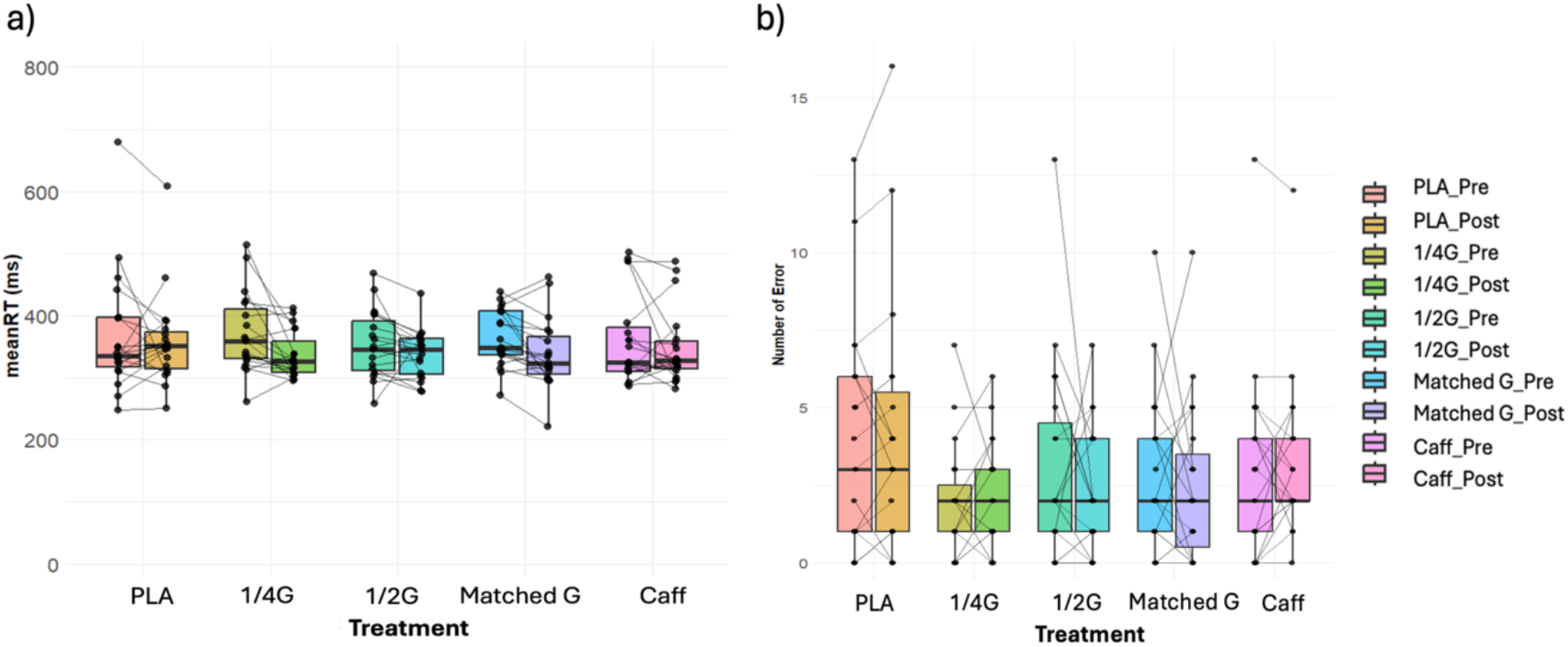
Effects of treatments on INH task performance with a) Mean Reaction Time and b) Number of Errors. Dots represent individual performance pre- and post-supplementation; lines connect repeated measures for each participant. RT = reaction time; PLA = placebo; Caff = caffeine; 1/4G, 1/2G, and Matched G represent guarana doses containing 1 mg⋅kg⋅bw^-1^, 2 mg⋅kg⋅bw^-1^, and 4 mg⋅kg⋅bw^-1^ caffeine.

In the INH measure, no significant main effects observed for treatment (*F*_(4,72)_ = 0.49, *p* = 0.66, η² = 0.01), time (*F*_(1,18)_ = 4.42, *p* = 0.05, η² = 0.02), or treatment×time interaction (*F*_(4,72)_ = 1.09, *p* = 0.36, η² = 0.01). This pattern was mirrored in error-related outcomes, where none of the comparisons reached statistical significance for either main effects of treatment (*F*_(1,18)_ = 2.32, *p* = 0.10, η² = 0.04), time (*F*_(1,18)_ = 0.06, *p* = 0.81, η² < 0.01), or interaction terms (*F*_(4,72)_ = 0.91, *p* = 0.43, η² < 0.01), as presented in *Supplementary Material 1*.

### 3.4 Word Memory Task Performance

Individual participant performance across timepoints and treatment conditions is visualised in *Figure 9*, illustrating a wide range of variability in memory task outcomes.

**Figure 9.**
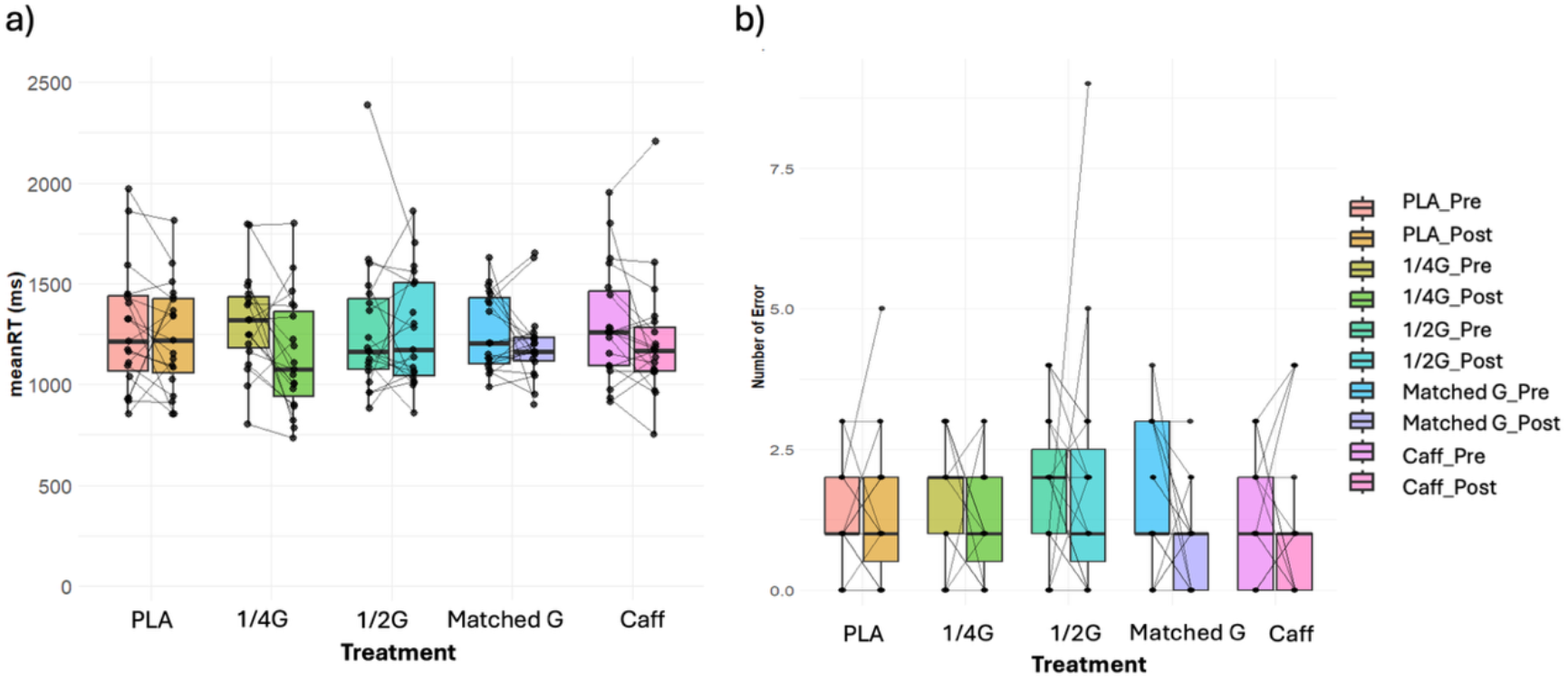
Effects of treatments on MEM task performance with a) Mean Reaction Time and b) Number of Errors. Dots represent individual performance pre- and post-supplementation; lines connect repeated measures for each participant. RT = reaction time; PLA = placebo; Caff = caffeine; 1/4G, 1/2G, and Matched G represent guarana doses containing 1 mg⋅kg⋅bw-1, 2 mg⋅kg⋅bw-1, and 4 mg⋅kg⋅bw-1 caffeine.

The same analysis was conducted with the memory task data, with the only significant effect of time was observed (*F*_(1,18)_ = 0.06, *p* = 0.81, η² < 0.01), indicating overall performance changes between pre- and post-ingestion. Further post-hoc comparisons revealed a significant reduction between timepoints (t (9) = 2.59, p = 0.05). However, no significant main effects observed for treatment type (*F*_(4,72)_ = 0.26, *p* = 0.87, η² < 0.01) and treatment×time interaction (*F*_(4,72)_ = 1.03, *p* = 0.39, η² = 0.01). Error counts did not differ significantly across treatments (*F*_(1,18)_ = 2.31, *p* = 0.09, η² = 0.04), time (*F*_(1,18)_ = 0.80, *p* = 0.38, η² < 0.01), and in treatment×time interaction (*F*_(4,72)_ = 0.93, *p* = 0.43, η² = 0.02), seen on *Supplementary Material 1*.

### 3.5 Effects of Supplements on Changes in Reaction Time

To account for the potential influence of covariates, including habitual caffeine intake, age, gender, and body fat percentage, multiple linear regression models were constructed to examine the isolated effects of treatments on changes in RT in all four cognitive tasks compared to the placebo, while controlling for confounding variables. Multiple linear regression models revealed no statistically significant effects for the SRT task (*F*_(8, 86)_ = 0.61, *p* = 0.77, *R²* _adjusted_ = −0.034), MEM task (*F*_(8, 86)_ = 0.98, *p* = 0.46, *R²* _adjusted_ = -0.002) or the SA task (*F*_(8, 86)_ = 0.59, *p* = 0.78, *R²* _adjusted_ = −0.036). Furthermore, none of the individual predictors significantly contributed to changes in RT for all 3 tasks (*p* > 0.05, shown in *Table 2*). However, a multiple linear regression predicting changes in RTs in inhibition task performance revealed a statistically significant overall model (*F*_(8,86)_ = 2.25, *p* = 0.03, *R²* _adjusted_ = 0.1).

**Table 2.** Multiple Linear Regression Results for Changes in Reaction Time on SRT, SA, INH and MEM Tasks.

| Determination | Characteristics | Est. Coeff | Std. Error | t | *p* |
|---|---|---|---|---|---|
| 1) Simple Reaction Time Task |  |  |  |  |  |
| Adjusted R<sup>2</sup> = -0.034 | (Intercept) | 21.18 | 62.30 | 0.34 | 0.74 |
| F(8, 86) = 0.61 | Treatment (1/4 Guarana) | 14.60 | 20.13 | 0.73 | 0.47 |
| *p* = 0.77 | Treatment (1/2 Guarana) | 5.30 | 20.15 | 0.26 | 0.79 |
|  | Treatment (Matched Guarana) | 28.78 | 20.13 | 1.43 | 0.16 |
|  | Treatment (Caffeine) | 19.49 | 20.12 | 0.97 | 0.34 |
|  | Daily Caffeine Intake (mg/day) | 0.13 | 0.14 | 0.93 | 0.36 |
|  | Age (years) | -2.03 | 2.17 | -0.94 | 0.35 |
|  | Gender | -10.99 | 21.79 | -0.51 | 0.62 |
|  | Body Fat Percentage (%) | 0.25 | 1.40 | 0.18 | 0.86 |
| 2) Sustained Attention Task |  |  |  |  |  |
| Adjusted R<sup>2</sup> = -0.036 | (Intercept) | 56.36 | 91.80 | 0.61 | 0.54 |
| F(8, 86) = 0.59 | Treatment (1/4 Guarana) | 39.29 | 29.67 | 1.33 | 0.19 |
| p = 0.78 | Treatment (1/2 Guarana) | 9.09 | 29.69 | 0.31 | 0.76 |
|  | Treatment (Matched Guarana) | 2.93 | 29.66 | 0.10 | 0.92 |
|  | Treatment (Caffeine) | 9.69 | 29.65 | 0.33 | 0.75 |
|  | Daily Caffeine Intake (mg/day) | 0.27 | 0.21 | 1.31 | 0.20 |
|  | Age (years) | -2.14 | 3.19 | -0.67 | 0.51 |
|  | Gender | -17.87 | 32.1 | -0.56 | 0.58 |
|  | Body Fat Percentage (%) | -0.93 | 2.06 | -0.45 | 0.65 |
| 3) Inhibition Task |  |  |  |  |  |
| Adjusted R<sup>2</sup> = 0.096 | (Intercept) | -14.82 | 52.84 | -0.28 | 0.78 |
| F(8, 86) = 2.25 | Treatment (1/4 Guarana) | 25.75 | 17.08 | 1.51 | 0.14 |
| p = 0.031 | Treatment (1/2 Guarana) | 6.51 | 17.09 | 0.38 | 0.70 |
|  | Treatment (Matched Guarana) | 17.94 | 17.07 | 1.05 | 0.30 |
|  | Treatment (Caffeine) | 5.50 | 17.07 | 0.32 | 0.75 |
|  | Daily Caffeine Intake (mg/day) | 0.13 | 0.12 | 1.11 | 0.27 |
|  | Age (years) | 0.57 | 1.84 | 0.31 | 0.76 |
|  | Gender | -25.09 | 18.48 | -1.36 | 0.18 |
|  | Body Fat Percentage (%) | 1.08 | 1.19 | 0.91 | 0.37 |
| 4) Memory Task |  |  |  |  |  |
| Adjusted R<sup>2</sup> = -0.0018 | (Intercept) | -276.88 | 263.99 | -1.05 | 0.30 |
| F(8, 86) =0.98 | Treatment (1/4 Guarana) | 137.91 | 85.31 | 1.62 | 0.11 |
| p = 0.46 | Treatment (1/2 Guarana) | -21.10 | 85.39 | -0.25 | 0.81 |
|  | Treatment (Matched Guarana) | 29.42 | 85.30 | 0.35 | 0.73 |
|  | Treatment (Caffeine) | 46.38 | 85.27 | 0.54 | 0.59 |
|  | Daily Caffeine Intake (mg/day) | 0.40 | 0.59 | 0.68 | 0.50 |
|  | Age (years) | 0.04 | 9.18 | 0.0050 | 0.99 |
|  | Gender | 169.61 | 92.31 | 1.84 | 0.070 |
|  | Body Fat Percentage (%) | 9.38 | 5.93 | 1.58 | 0.12 |

*Note: RT = reaction time; PLA = placebo; Caff = caffeine; 1/4G, 1/2G, and Matched G represent guarana doses equivalent to 1 mg/kg, 2 mg/kg, and 4 mg/kg BW caffeine.*

## 4. Discussion

This study is among the first to examine the dose-response effects of guarana and its direct comparison with caffeine on four cognitive domains: alertness, concentration, inhibitory control, and short-term memory. The findings demonstrated that acute guarana supplementation did not significantly improve these cognitive domains compared to placebo. The observed effects in SRT, SA, and INH were minimal and inconsistent, while results from MEM further supported the absence of any acute cognitive enhancement from either supplement. Furthermore, no significant differences were observed between guarana and a matched caffeine dose in these four cognitive domains, suggesting that caffeine does not exert additional cognitive benefits beyond those provided by guarana.

While the results of the current study did not reach statistical significance, the findings suggest that guarana may offer cognitive benefits comparable to those of caffeine, even at lower equivalent doses (e.g., half and quarter doses). Given its natural origin and the presence of additional bioactive compounds, guarana could serve as a viable alternative for individuals seeking plant-based or lower-risk cognitive enhancers. Future product development and public health guidance may consider incorporating guarana as a functional ingredient, especially for populations aiming to reduce caffeine intake without compromising cognitive performance.

The non-significant improvements in cognitive performance following guarana supplementation, align with previous findings. Pomportes et al. (17) reported non-significant effects of a vitamin/mineral/guarana complex (containing 300 mg guarana) and a matched 100mg caffeine supplementation on mean RT during SRT in 56 healthy adults. Among other studies that closely aligned with ours, Talik et al. (23) reported similarly null effects using a randomized, double blind, within-subjects design. In their sample of 40 healthy adults, participants consumed 3 mg/kg of guarana and completed Go/No-Go and 2N-back tasks (23). Given the comparable sample characteristics, dosing strategy and validated tasks, their findings reinforce our conclusion that acute guarana supplementation has limited cognitive benefits in non-fatigued conditions. Similarly, Kennedy et al. (15) found no significant improvements in either accuracy in a digit vigilance task, which assessed concentration, or mean RT in SRT following administration of capsules containing 75 mg guarana powder to 28 young adults. However, that study also found significantly faster RTs at 1 hour (-16.1 ms), 4 hours (-15.0 ms), and 6 hours (-26.6 ms) post-dose in the digit vigilance task (15), suggesting the presence of time-dependent effects. In contrast, the current study only assessed post-supplementation performance 30 minutes after ingestion. Studies have shown that caffeine reaches peak plasma concentrations between 30-60 minutes post-ingestion, with 99% absorption occurring by 45 minutes (30), suggesting that a longer interval may have allowed for greater absorption and potentially stronger cognitive effects. Furthermore, Gurney et al. (14) observed a 17 ms reduction in mean RT in SRT following 125 mg⋅kg⋅bw^-1^ guarana after a V̇O_2_max test, but no improvements in accuracy were observed pre- or post-exercise in 27 physically active participants. The observed improvement in RT is more likely contributed by exercise-induced effects rather than guarana alone. Exercise has been shown to decrease RT by reducing motor response latency through the optimisation of muscular tension (44), increasing cerebral blood flow, and modulating the production of neurotransmitters such as dopamine (45,46).

The present study did not show a dose-response relationship between guarana and improvements in cognitive performance, aligning with a meta-analysis that reported no significant association between the guarana dose and the effect size of cognitive benefits across studies (22). However, these results contradict Haskell et al. (17), who tested 26 participants consuming 37.5 mg, 75 mg, 150 mg, and 300 mg guarana capsules on four separate days. They found that RTs in digit vigilance and memory tasks improved most with the lower doses (37.5 mg, 75 mg), while subjective alertness, assessed by the Bond–Lader mood scale, was only enhanced at the highest dose (300 mg) (16). Despite some observed trends, no statistically significant improvements were observed in attentional task performance, and as Haskell’s study assessed alertness subjectively, outcomes may have been influenced by perceptual bias. Amaritei et al (47) also reported similar perceived cognitive improvements in testing visual memory and learning scores following 10 days of guarana (2 g⋅day^-1^) combined with green tea (1.5 g⋅day^-1^) in healthy students. However, the open-label design, subjective outcomes, and inability to isolate guarana’s effects limit the applicability of those findings to objective performance outcomes (47). Moreover, differences in dosing strategies may explain the divergence in findings. Unlike Haskell et al. and Amaritei et al’s studies, which employed fixed-dose administration, the current study utilised bodyweight-adjusted dosing. As a result, participants in the present study received substantially higher absolute guarana doses, which may have elicited different pharmacodynamic responses compared to fixed-dose trials. Pomportes et al. (17) reported participants exhibited significantly faster reaction times on the Go/No-Go task at 30-, 60-, and 90-minutes post-ingestion of the guarana complex compared to caffeine (17). These cognitive benefits were accompanied by increased parasympathetic activity (21.2 ± 2.6%) indexed by heart rate variability (HRV), suggesting a physiological mechanism supporting enhanced attentional regulation and higher cognitive functions. Furthermore, Haskell et al. (16) and Kennedy et al. (15) reported cognitive enhancements in alertness and concentration by guarana doses containing 4.5 mg (0.06 mg⋅kg⋅bw^-1^ for a 75kg individual) and 9 mg (0.12 mg⋅kg⋅bw^-1^) caffeine, both below the pharmacological caffeine threshold (0.5 mg⋅kg⋅bw^-1^) (48). In both studies, improvements were observed in tasks requiring sustained attention and working memory (15,16). These results lend further support to indicate that other bioactive components in guarana may act synergistically with caffeine to enhance cognitive performance (49).

Several mechanisms have been proposed through which non-caffeine bioactive compounds in guarana may enhance cognitive function. Theobromine, a caffeine derivative, acts as a non-selective adenosine receptor antagonist and a phosphodiesterase inhibitor, increasing intracellular cyclic adenosine monophosphate (cAMP) levels (50). This activates downstream signalling cascades to increase brain-derived neurotropic factor (BDNF) production, promoting neurogenesis and synaptic strength (51). However, theobromine is less potent than caffeine but has a longer half-life, contributing to prolonged or delayed arousal (49). Guarana is also rich in polyphenols, including tannins and catechins (1).

Tannins have been shown to reduce lipid peroxidation and help to preserve endogenous antioxidant activity, thereby reducing oxidative stress in the brain (52). They also attenuate neuroinflammation by downregulating proinflammatory cytokines (IL-6, IL-1β, TNF-α) via suppression of the NF-κB pathway (53) and modulate apoptosis and autophagic pathway to support cognitive health (54). Similarly, catechins have shown to exhibit antioxidant and neuroprotective effects that potentially benefit chronic brain conditions (55). Whereas the neurocognitive impact of these compounds likely requires sustained exposure or specific stress contexts to manifest, our protocol involved brief, low cognitive-load tasks under rested conditions, which may not sufficiently activate the neural circuits modulated by these slower-acting phytochemicals. Moreover, compounds such as tannins may form complexes with caffeine via non-covalent interactions, such as hydrogen bonding and aromatic-aromatic interactions (56,57), potentially extending caffeine’s half-life (58) but reducing its absorption rate to delay its stimulant effects (59). Since the bioavailability and temporal kinetics of bioactive compounds in guarana are not well characterised in combination (1), it can possibly lead to suboptimal timing of cognitive benefit during acute testing windows. Future research should investigate guarana’s time-dependent effects and further elucidate the mechanisms of its active constituents.

This study acknowledges several limitations. Firstly, the four tasks do not represent the full spectrum of cognitive functions relevant to daily life, therefore future studies should incorporate additional cognitive tasks examining other cognitive functions, such as decision-making (60), for a more comprehensive evaluation of cognitive function. Secondly, the simplicity of tasks used may result in ceiling effects, as high initial accuracy is known to limit sensitivity to post-intervention changes (61), which can account for the absence of significant findings. While session-based improvements in reaction time and memory were observed, these gains occurred uniformly across all conditions, as well as being statistically non-significant, likely reflecting practice effects. The absence of pharmacokinetic data from the guarana doses and caffeine in our participants prevents confirmation of temporal effects of interventions on central activation, which is crucial for interpreting psychostimulant studies (62). Testing was conducted under non-fatigued conditions, which may have dampened the expression of guarana’s cognitive effects, as previous studies have shown greater stimulant responsiveness under stress or cognitive load (7,14,19). Additionally, our participant cohort were primarily young healthy adults which limits the generalisability of the findings as the cognitive benefits of guarana’s constituents have been observed in ageing and clinical populations, including individuals with cognitive diseases such as Alzheimer’s (63,64), highlighting the need for further research to assess guarana’s effects within these groups. It is also worth mentioning that cognitive performance can be affected by the body’s natural circadian and individual chronotypes (65). Although many participants were tested at consistent times, several completed assessments at varying time points, introducing potential time-of-day effects. Lastly, though a double-blind design was employed, guarana’s distinctive taste and texture may have compromised participant blinding. The exit questionnaire indicated a 30% correct identification rate, suggesting a high likelihood of the presentation of participant bias. To combat this, providing guarana in capsules may improve blinding, but the influence of ingestion matrices on cognitive outcomes needs further investigation.

## 5. Conclusion

This study found no statistically significant dose-response effects of acute guarana supplementation or any differences compared to caffeine over time on cognitive performance, including alertness, concentration, inhibitory control, and memory, in healthy adults. Nevertheless, the comparable effects of guarana to caffeine at all tested doses highlight its potential as a natural, plant-based alternative for individuals seeking to reduce caffeine intake. Given its unique phytochemical profile, guarana may exert more pronounced effects under conditions of stress, fatigue, or with chronic supplementation. Future research should explore these contexts and incorporate physiological and neurochemical biomarkers. Also, a better understanding of guarana’s underlying mechanisms is essential to determine its suitability for cognitive enhancement in both clinical and real-world settings.

## Author Contribution

TD, TG, ZH, and TL conceived and designed the experiments; ZH and TL performed the experiments; ZH and TL analysed the data; ZH and TL wrote the paper.

## Financial Support

This research received no specific grant from any funding agency, commercial or not-for-profit sectors.

## Declaration of interests

The authors declare none.

## Data Availability

All data produced in the present study are available upon reasonable request to the authors

## Data Availability Statement

The data supporting the findings of this study are available from the corresponding author upon reasonable request.

## List of Abbreviations

BW: Body Weight
INH: Inhibition
MEM: Memory
SA: Sustained Attention
SRT: Simple Reaction Time

## Notes

### Competing Interest Statement

The authors have declared no competing interest.

### Author Declarations

The Research Ethics Committee of University College London gave ethical approval for this work

