## Supplementary Table S1 for "Acute Dose-Response Effects of Guarana and Caffeine on Cognitive Performance in Healthy Adults"

**Supplementary Table: Cognitive Task Performance Across Treatments and Times**

**Table S1 Mean ± SD values for RT and number of errors in simple reaction time, sustained attention, inhibition and memory tasks before and after supplementation across all treatment conditions.**

*RT = reaction time; PLA = placebo; Caff = caffeine; 1/4G, 1/2G, and Matched G represent guarana doses equivalent to 1 mg/kg, 2 mg/kg, and 4 mg/kg BW caffeine).*

| **Test** | **Variable** | **Treatment** | **Pre-supplementation** | **Post-supplementation** |
| --- | --- | --- | --- | --- |
| Simple Reaction Time | Mean RT (ms) | PLA | 304.05 ± 40.52 | 323.27 ± 53.33 |
|  |  | 1/4G | 302.37 ± 40.80 | 307.10 ± 55.58 |
|  |  | 1/2G | 298.41 ± 38.57 | 312.52 ± 59.52 |
|  |  | Matched G | 323.45 ± 76.45 | 314.00 ± 63.24 |
|  |  | Caff | 309.16 ± 37.25 | 308.89 ± 43.03 |
|  | Number of Errors | PLA | 0.47 ± 0.84 | 0.68 ± 0.75 |
|  |  | 1/4G | 0.58 ± 0.90 | 0.47 ± 0.61 |
|  |  | 1/2G | 0.79 ± 0.92 | 0.47 ± 0.61 |
|  |  | Matched G | 0.58 ± 0.84 | 0.42 ± 0.84 |
|  |  | Caff | 0.42 ± 0.84 | 0.53 ± 0.70 |
| Sustained Attention | Mean RT (ms) | PLA | 575.51 ± 176.20 | 579.90 ± 181.58 |
|  |  | 1/4G | 534.25 ± 127.02 | 498.91 ± 93.15 |
|  |  | 1/2G | 525.44 ± 103.06 | 520.02 ± 136.60 |
|  |  | Matched G | 556.61 ± 168.35 | 557.71 ± 206.53 |
|  |  | Caff | 544.15 ± 104.44 | 538.84 ± 119.24 |
|  | Number of Errors | PLA | 0.42 ± 0.61 | 0.37 ± 0.76 |
|  |  | 1/4G | 0.53 ± 0.61 | 0.42 ± 0.69 |
|  |  | 1/2G | 0.26 ± 0.45 | 0.26 ± 0.45 |
|  |  | Matched G | 0.47 ± 0.61 | 0.11 ± 0.32 |
|  |  | Caff | 0.21 ± 0.43 | 0.21 ± 0.42 |
| Inhibition Task | Mean RT (ms) | PLA | 367.46 ± 98.38 | 358.84 ± 75.43 |
|  |  | 1/4G | 370.40 ± 64.30 | 336.53 ± 37.67 |
|  |  | 1/2G | 352.78 ± 53.79 | 338.48 ± 39.24 |
|  |  | Matched G | 364.32 ± 46.39 | 338.17 ± 56.60 |
|  |  | Caff | 362.16 ± 74.34 | 348.06 ± 60.41 |
|  | Number of Errors | PLA | 3.84 ± 3.82 | 4.11 ± 4.19 |
|  |  | 1/4G | 2.05 ± 1.78 | 2.26 ±1.66 |
|  |  | 1/2G | 3.05 ± 3.27 | 2.26 ± 2.02 |
|  |  | Matched G | 2.90 ± 2.58 | 2.47 ± 2.61 |
|  |  | Caff | 2.84 ± 3.13 | 3.21 ± 2.62 |
| Memory Task | Mean RT (ms) | PLA | 1274.91 ± 309.41 | 1238.09 ± 267.70 |
|  |  | 1/4G | 1313.42 ± 246.40 | 1142.99 ± 288.32 |
|  |  | 1/2G | 1276.10 ± 345.53 | 1267.54 ± 281.23 |
|  |  | Matched G | 1258.00 ± 193.30 | 1195.32 ± 185.83 |
|  |  | Caff | 1298.92 ± 287.53 | 1215.82 ± 305.40 |
|  | Number of Errors | PLA | 1.32 ± 0.89 | 1.42 ± 1.30 |
|  |  | 1/4G | 1.63 ± 1.01 | 1.21 ± 0.98 |
|  |  | 1/2G | 1.79 ± 1.32 | 1.89 ± 2.18 |
|  |  | Matched G | 1.58 ±1.26 | 0.89 ± 0.81 |
|  |  | Caff | 1.16 ± 1.12 | 1.16 ± 1.38 |
